# An Explainable Cryobiopsy AI Model, CRAI, to Predict Progression in Interstitial Pneumonia

**DOI:** 10.1101/2025.08.06.25333181

**Authors:** Wataru Uegami, Ethan N. Okoshi, Kris Lami, Yuichiro Nei, Mutsumi Ozasa, Kensuke Kataoka, Yuka Kitamura, Yasuo Kohashi, Lee A.D. Cooper, Hidenori Sakanashi, Yuji Saito, Yasuhiro Kondoh, the study group on CRYOSOLUTION, Junya Fukuoka

## Abstract

Interstitial lung disease (ILD) encompasses diverse pulmonary disorders with varied prognoses. Current pathological diagnoses suffer from inter-observer variability, necessitating more standardized approaches. We developed an ensemble AI model for cryobiopsy, CRAI, an artificial intelligence model to analyse transbronchial lung cryobiopsy (TBLC) specimens and predict patient outcomes. We developed an explainable AI model, CRAI, to analyse TBLC. CRAI comprises seven modules for detecting histological features, generating 17 pathologically significant findings. A downstream XGBoost classifier was developed to predict disease progression using these findings. The model’s performance was evaluated using respiratory function changes and survival analysis in cross-validation and external test cohorts. In the internal cross-validation (135 cases), the model predicted 105 cases without disease progression and 30 with disease progression. The annual Δ%FVC was -1.293 in the non-progressive group versus -5.198 in the progressive group, outperforming most pathologists’ diagnoses. In the external test cohort (48 cases), the model predicted 38 non-progressive and 10 progressive cases. Survival analysis demonstrated significantly shorter survival times in the progressive group (*p* = 0.034). CRAI provides a comprehensive, interpretable approach to analysing TBLC specimens, offering potential for standardizing ILD diagnosis and predicting disease progression. The model could facilitate early identification of progressive cases and guide personalized therapeutic interventions.

## 1 Introduction

Interstitial lung disease (ILD) is a diverse group of pulmonary disorders that vary in prognosis and treatment approaches.^1^ For instance, idiopathic pulmonary fibrosis (IPF) is a progressive condition with a poor prognosis, often treated by anti-fibrotic medications.^2^ In contrast, non-specific interstitial pneumonia (NSIP), hypersensitivity pneumonitis^3^ (HP), and connective tissue disease-associated ILDs^4^ (CTD-ILDs) typically require treatment targeting inflammatory changes or the underlying causes, such as inhaled antigens or autoimmune conditions. Therefore, accurate diagnosis is crucial for determining appropriate treatment strategies and predicting patient outcomes.

Recent advancements have revealed the presence of poor prognostic subgroups within non-IPF interstitial lung diseases.^1, 5^ The latest IPF guideline introduced the concept of progressive pulmonary fibrosis (PPF), which is defined as at least two of the following criteria: worsening respiratory symptoms, physiological evidence of disease progression, and radiological evidence of disease progression occurring within the past year in patients with ILDs other than IPF.^1^ Additionally, multiple clinical trials have tested anti-fibrotic drugs as a treatment for various progressive ILDs.^6–8^ There is growing attention on methods to identify progressive cases early to perform therapeutic interventions at the appropriate time.

The current concept of PPF is inherently retrospective, as it requires evidence of clinical, physiological, or radiological worsening over time. Patients can only be recognized as having progressive disease after progression has already occurred. If future disease progression could be predicted from pathological findings at the time of biopsy, high-risk patients could be identified earlier, potentially allowing timely therapeutic intervention before irreversible functional decline develops; however, histological studies on progressive cases are still few and were not included in the 2022 criteria for PPF. Tsushima et al. have reported that the focal usual interstitial pneumonia (UIP) pattern is a key prognostic pattern in PPF.^9^ UIP is characterized by temporally and spatially heterogeneous fibrosis, manifesting as a patchy distribution of immature and dense fibrotic areas.^1, 10^ This pattern reflects the progressive nature of the disease and has significant implications for patient prognosis and treatment decisions. Moreover, the reliability of pathological evaluations has been challenged by low inter-observer agreement among pathologists.^11, 12^ This inconsistency raises significant concerns, as it can lead to misdiagnoses, resulting in patients receiving inappropriate treatments or missing out on beneficial therapies, which may accelerate disease progression.

The advent of artificial intelligence (AI)-based image analysis offers promising potential for standardizing pathological diagnoses and addressing the challenge of inter-observer variability.^13, 14^ Our previous work introduced MIXTURE, the first and only reliable AI model designed to analyze surgical lung biopsy (SLB) specimens in interstitial lung disease.^15^ MIXTURE demonstrated high accuracy in detecting UIP patterns in SLB samples, with patients classified as UIP by the model showing significantly poorer survival outcomes.

While SLB has traditionally been the gold standard for the pathological diagnosis of ILD, its invasive nature^16^ has limited its application. In contrast, the less invasive transbronchial lung cryobiopsy (TBLC) has gained prominence.^1, 17^ However, SLB and TBLC specimens differ substantially in both qualitative and quantitative aspects, presenting unique challenges for AI analysis.

In this study, we present CRAI for cryobiopsy, a novel ensemble AI model specifically designed to predict future disease progression from TBLC specimens obtained at the time of diagnosis. By extracting pathologically interpretable histological features from H&E-stained sections, CRAI aims to identify patients at high risk of progression before clinical deterioration becomes evident. This approach may support earlier therapeutic decision-making and provide a pathological basis for precision management of ILD.

## 2 Methods

### 2.1 Patients and Datasets

Five datasets were constructed from the two institutions (Table 1). The TGH-SLB set was collected at Tosei General Hospital (TGH) and consulted at Nagasaki University, containing pathological whole slide images (H&E and EVG staining) of SLB cases, sampled between 2015 and 2020. This dataset was used for model training to supplement the limited number of TBLC cases. The TGH-TBLC set, used for model training and validation, consisted of TBLC cases collected at Tosei General Hospital and consulted at Nagasaki University, sampled between 2019 and 2023. The inclusion criteria was patients whom TBLC and any follow-up pulmonary function tests (PFTs) were available. This dataset included histopathological slides, pulmonary function test (PFT) data (% forced vital capacity, %FVC), and survival data. The baseline PFT was the most recent test performed less than 100 days prior to the TBLC procedure, and follow-up PFTs at one and two years were also collected when available. The HRH-PFT and HRH-survival set, used as a held-out external test set, included consecutive TBLC cases diagnosed at Haruhi Respiratory Medical Hospital (HRH) for PFT analysis and survival analysis respectively. All pathological specimens above were scanned using an Aperio CS2 whole slide image (WSI) scanner at 20× or 40× magnification. The KMC set consisted of consecutive TBLC cases collected at Kameda Medical Center between 2019 and 2024, with at least two year of clinical follow-up. Pathological specimens were scanned at 40× magnification using a Philips Ultra Fast Scanner and exported as TIFF files. This research plan was approved by the Ethics Committee of Nagasaki University Hospital (approval number: 19081929-4). The requirement for informed consent was waived due to the retrospective nature of the study.

**Table 1:** Summary of cohort characteristics and baseline details. Data are n, n (%) or mean±SD. PFT=pulmonary function test, SLB=surgical lung biopsy, TBLC=transbronchial lung cryobiopsy, TBLB=transbronchial lung biopsy, FVC=forced vital capacity, *DL*_*CO*_=Diffusing capacity for carbon monoxide, prog=progression. Disease progression was defined as ≥5% absolute decline in percentage for each year over a two-year observation period.

|  | TGH-SLB | TGH-TBLC | HRH-PFT | HRH-survival | KMC |
| --- | --- | --- | --- | --- | --- |
| Purpose | Model development | Cross validation | External test PFT analysis | External test survival analysis | External test |
| Modality | SLB, TBLC, TBLB | TBLC | TBLC | TBLC | TBLC |
| No. of cases | 53 | 135 | 30 | 48 | 42 |
| Scanner | Aperio CS2 | Aperio CS2 | Aperio CS2 | Aperio CS2 | Philips UFS |
| File format | svs | svs | svs | svs | tiff |
| Male sex | 31 (58.5) | 70 (51.5) | 18 (36) | 28 (58) | 31 (74) |
| Age (yr) | 59.57±11.91 | 66.6±9.78 | 70.2±8.4 | 66.7±14.0 | 70.9±8.65 |
| Former/ current smoker | - | 8 (26.7) | 16 (33.3) | - | - |
| PFT |  |  |  |  |  |
| %FVC, baseline | - | 92.8±20.4 | 89.9±18.6 | 90.3±20.9 | 91.2±21.79 |
| %$DL_{CO}$, baseline | - | 72.7±22.4 | 74.5±23.0 | 71.8±26.7 | - |
| Disease progression |  |  |  |  |  |
| Prog-Prog | - | 7 (5.2) | - | - | - |
| Prog-Non prog | - | 12 (8.9) | - | - | - |
| Non prog-Prog | - | 30 (22.2) | - | - | - |
| Non prog-Non prog | - | 86 (63.7) | - | - | - |
| Treatment |  |  |  |  |  |
| Antifibrotic agents | - | 35 (25.9) | - | - | - |
| Pirfenidone | - | 8 (5.9) | - | - | - |
| Nintedanib | - | 29 (21.5) | - | - | - |
| Anti-inflammatory | - | 65 (48.1) | - | - | - |
| Oral | - | 63 (46.7) | - | - | - |
| Steroid infusion | - | 45 (33.3) | - | - | - |

### 2.2 Rationale for Outcome Selection

Disease progression was selected as the primary predictive outcome because it represents a clinically relevant endpoint in ILDs, closely associated with functional decline and mortality across diagnostic categories. Given the substantial inter-observer variability in TBLC-based pathology, progression provides an objective and clinically actionable target for risk stratification and treatment planning.

### 2.3 Model Overview

CRAI consists of seven different modules that detect basic histological structures (lung-stroma model, 1× model, 5× and 20× models, elastic and collagen fiber models, and lymphocyte model), detailed in the supplementary material. By integrating these structures at the slide level, 17 pathologically significant slide-level features are calculated. Finally, a downstream XGBoost classifier is constructed using these 17 findings as input to predict patients’ disease progression (Figure 1). The models are summarized in Table S1 and the examples of each models’ outputs are presented in Figure 2 and S3. No data preprocessing was performed for the models construction.

**Figure 1:**
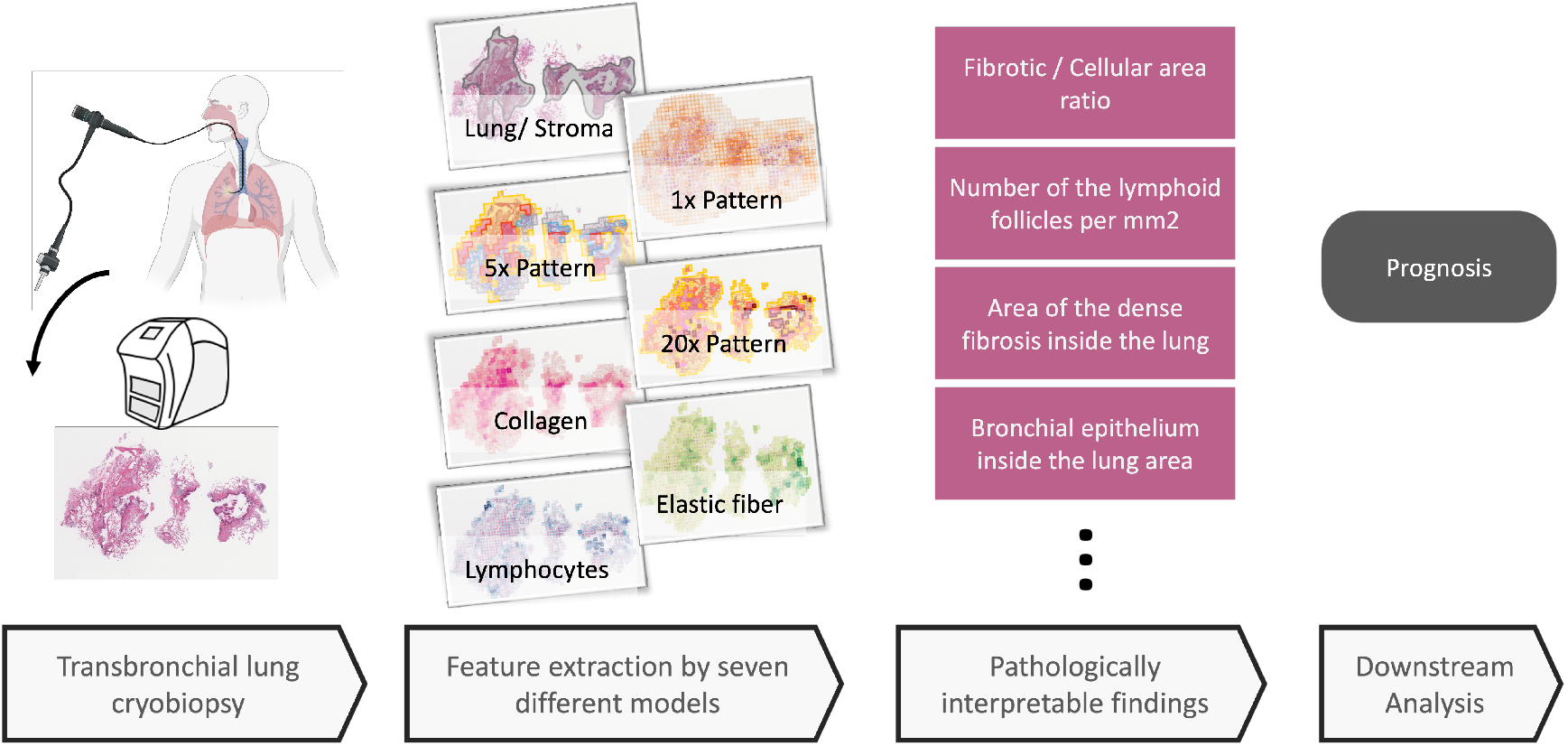
Workflow of CRAI: A Multi-model Approach for Prognostic Prediction. Transbronchial lung cryobiopsy specimens undergo comprehensive analysis through seven specialized deep learning models: the lung-stroma model, classification models optimized for 1×, 5×, and 20× magnifications, and models for collagen, elastica, and lymphocyte detection. Each model extracts distinct pathological features, which are then consolidated to generate 17 pathologically interpretable slide-level findings. These parameters are subsequently integrated into an XGBoost-based downstream model for disease progression prediction. Created in https://BioRender.com

**Figure 2:**
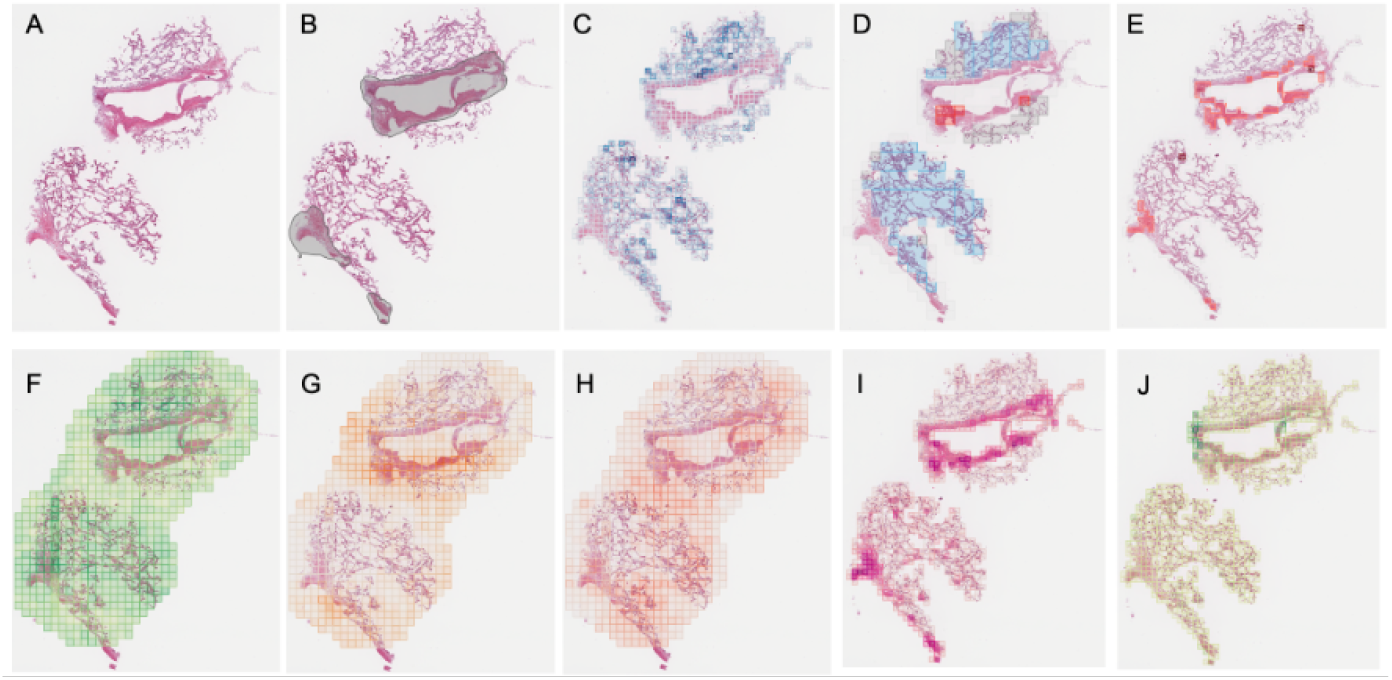
Representative results of CRAI. (A) Original H&E-stained whole slide image. (B) Lung-stroma model highlighting broncovascular bundles in gray. (C) Spatial distribution of lymphocyte density as predicted by the lymphocyte model. (D) Classification results of the 5× model, where *fibrotic areas* are shown in red, *cellular areas* in light blue, *lymphoid follicles* in blue (not present in this case), *normal tissue* in gray, and *other findings* in white. (E) Results of the 20× model, with *fibrosis* marked in orange, *bronchiolar epithelium* in brown, *FP/OP pattern* in pink, mucin in yellow, and *other findings* in gray. (F-H) Predictions of three separate 1× models, showing the distribution of *diffuse, patchy*, and *honeycomb* patterns, respectively. (I) Predicted distribution of collagen fibers by the collagen model. (J) Predicted distribution of elastic fibers by the elastica model.

### 2.4 Lung-Stroma Model

This model is designed to distinguish the bronchovascular bundle and lung parenchymal regions, which are relatively abundant in TBLC specimens. The model was implemented as a segmentation model for bronchovascular bundles in the specimen. Annotations were provided for 45 TBLC whole slide images, and an EfficientNet-b0^18^backbone U-Net^19^ was trained on 512px (3mm) square patches.

### 2.5 1× Model

This model is designed to detect the distribution of three distinct tissue patterns: *honeycomb, patchy*, and *diffuse*, which can be recognized at low magnification (1.25×). A total of 192 slides (72 cases) diagnosed with UIP from the TGH-SLB dataset were utilized as training data. The patch size was set to 460 pixels (3.68mm) square. A pathologist specializing in ILD classified the patches as *patchy* using a web application.^20^ Patches classified as *patchy* were defined as those containing a mixture of lesions and normal lung tissue. In a similar manner, patches identified as *honeycomb* were selected from 72 slides (57 cases) diagnosed with UIP, where the honeycomb pattern is characterized by cystic spaces surrounded by dense fibrosis. Conversely, patches considered to be *diffuse* were selected from 192 slides (71 cases) diagnosed as Non-UIP, with the diffuse pattern defined as patches where lesions occupy the area without any normal lung tissue. This classification resulted in 2,268, 1,617, and 1,704 unique patches for the honeycomb, patchy, and diffuse categories, respectively. Common features of the patches were extracted using KimiaNet,^21^ and a multi-layer perceptron (MLP) regressor head was trained to calculate the probability of each feature’s presence. Optimal hyperparameters were determined through grid search with 5-fold cross-validation.

### 2.6 5× and 20× Models

These multi-class classification models classify tissue patterns at a moderate (5×) and high magnification (20×). It utilizes MIXTURE, a previously published model design from our earlier work^15^. To avoid semantic overlap with the newly developed model, some classes were merged as shown in Table S2 and S3.

### 2.7 Collagen and Elastic Fiber Models

These models quantitatively estimate the deposition of two distinctive fibers, collagen and elastic fibers, from H&E images in cases of ILD. For the TGH-SLB set, registration of H&E and EVG specimens was performed using Deep Feature Based Registration.^22^ The implementation was modified from the TIA toolkit code.^23^ The registration results were reviewed, and 37 out of 150 successful WSIs were selected. A pathologist performed color deconvolution on these 37 EVG specimens using QuPath,^24^ separating collagen fibers (red) from elastic fibers (black). The localization information of these fibers was saved as 1*/*8 size PNG mask images. Based on this, corresponding patches of H&E specimens of the same size as the 20× model were matched with patches of EVG specimens and the resulting color deconvolution outputs for collagen and elastica masks. The deposition levels of collagen and elastic fibers in the H&E images were defined as the ratio of the mask areas of the corresponding collagen and elastica masks, and a regressor model was constructed to infer the deposition levels of each fiber. The model architecture was ResNet18 pre-trained on ImageNet.

### 2.8 Lymphocyte Model

This model detects lymphocytes. Inflammatory cells were identified using HoverNet^25^ with weights from the PanNuke dataset^26^ resulting in 194,881 segmentation instances. A YOLOv7-based detection model was then constructed for rapid identification of inflammatory cells in 20× images.

### 2.9 Slide Level Integration

From the features identified by each model and their combinations, the following 18 findings were obtained: (1) lung, (2) lesion, (3) fibrotic/cellular ratio, (4) lymphoid follicles, (5) dense fibrosis, (6) bronchial epithelium, (7) number of fibroblastic proliferation/organizing pneumonia (FP/OP), (8) maximum collagen around the FP/OP, (9) lymphocyte density in dense fibrosis area, (10) lymphocyte density in non-dense fibrosis, (11) collagen in non-dense fibrosis area, (12) collagen edge, (13) collagen edge/dense fibrosis, (14) collagen forming the blob, (15) elastic fiber, (16) diffuse, (17) patchy, (18) honeycomb. The definition for each finding is shown in the supplementary materials. Cases where the lung area was estimated to be less than 10*mm*^2^ were excluded due to insufficient evaluable regions. This process requires no user intervention, as the model autonomously assesses tissue adequacy and suppresses predictions for insufficient TBLC specimens. All features other than lung were adopted as slide-level findings.

### 2.10 Construction of the Downstream Model for the Disease Progression Prediction

From the TGH-TBLC cohort, 135 cases with follow-up respiratory function data over 2 years were extracted. No missing data was reported. Progressive disease was defined as a decline of 5% or more in %FVC or a decline of 10% or more in percentage of the diffusing capacity of the lungs for carbon monoxide (%*DL*_*CO*_) within any single year, accompanied by no improvement in both functional tests from the baseline after two years of follow-up. The remaining cases were classified as the non-progressive group and served as labels for the training data; these served as labels for the training data. Using the 17 features extracted by AI as input, an XGBoost classifier was constructed to classify prognosis. XGBoost was selected based on our previous methodological experience.^27^ Hyperparameters were optimized through ten-fold cross-validation and grid search. Thus, for the model with the best hyperparameters, ten XGBoost models were trained, one for each cross-validation split. The model produced a binary output: progressive and non-progressive disease. For the inference values of the TGH-TBLC cohort, the inference results were synthesized from the cross-validation results. For external validation using the HRH-PFT cohort, the mode of the outputs from the above ten models was adopted as the output.

### 2.11 UIP Diagnosis by Multiple Pathologists

28 pathologtists independently observed 135 TBLC specimens, the same as above (TGH-TBLC). Referring to the WSIs, they classified the UIP pattern, which is well known as a histopathological prognostic factor. UIP patterns were categorized into two groups: UIP (present throughout the sample or occupying the majority of the lesion) and non-UIP (occupying a minor area of the lesion or completely absent). Analysis was performed using data from the 19 pathologists who diagnosed 130 or more cases.

### 2.12 Evaluation of CRAI and Pathologists’ UIP to Predict Δ%FVC

Patients with multiple recorded PFT measurements were selected for the study. The absolute change in %FVC (Δ%FVC) is considered an important predictor of mortality.^28^ For each pathologist, we applied a linear mixed model which included fixed effects for age, sex, and baseline %FVC, and a random effect for the interaction between UIP diagnosis and time (in years) to evaluate the differences in Δ%FVC by diagnosis group. To compare the pathologists’ diagnoses with CRAI, the same analysis was conducted using the output of CRAI (non-progressive vs. progressive group) instead of the UIP diagnosis. Statistical significance was determined using a 95% confidence interval.

### 2.13 Survival Analysis

This analysis was performed on the HRH-survival cohort. Overall survival was defined as the time from the date of clinical diagnosis to the date of death from any cause or the end of follow-up. Survival estimates were calculated using the Kaplan–Meier method for patient subgroups classified by AI, and these estimates were compared using the log-rank test. The level of significance was established *α* = 0.05.

### 2.14 Subset Analysis for Interstitial Lung Abnormality (ILA)

Test cases from TGH included cases categorized as interstitial lung abnormality based on clinical criteria.^29^ A subset analysis was conducted separately to evaluate prediction of progression in these cases.

### 2.15 Validation of the AI Model in an Independent Cross-Domain Cohort

The performance of the AI model for predicting disease progression was evaluated in an independent KMC cohort consisting of 42 cases. These cases were derived from a different institutional and imaging domain, including slides scanned with a different whole-slide scanner and prepared using independent staining protocols. Disease progression was defined as a decline of 5% or more in %FVC on pulmonary function testing within any single year. In this validation cohort, 23 cases showed disease progression and 19 cases remained non-progressive during follow-up.

### 2.16 Environment

The deep learning environment used Python 3.8.19 and Torch 1.13.1. Scikit-learn 1.3.0 and xgboost 2.1.1 was used for constructing the XGBoost models, and statsmodels 0.14.1 was used for the generalized linear mixed model analysis. Survival analysis was performed in the R 4.4.1 environment. The log-rank test used the survival package 3.6, and the survminer package 0.5.0 was used for plotting the Kaplan-Meier curves.

## 3 Results

### 3.1 Predictive Performance for Respiratory Function Deterioration

In the cross-validation of the TGH-TBLC cohort, CRAI predicted 105 cases as a non-progressive group and 30 cases as a progressive group. In the non-progressive group, the estimated annual Δ%FVC was -1.293, whereas in the progressive group, it was -5.198, with a statistically significant difference between the two groups (Figure 3, Table S4). Thirty patients were clinically categorized as having ILA, with ten of them identified as having a disease progression. The estimated Δ%FVC exhibited a similar trend (−2.853 in the non-progressive group and -8.093 in the progressive group, Table S5). Among the cases of ILD, some examples of the cases that were classified as progressive group by the model and actually showed clinically worsening respiratory function are shown in Figure S1. In external validation using the HRH-PFT cohort, 21 cases were inferred as a non-progressive and nine cases a progressive. The Δ%FVC in the non-progressive and progressive groups was -1.926 and -5.894, respectively, closely reflecting the results of the cross-validation cohort (Table S6).

**Figure 3:**
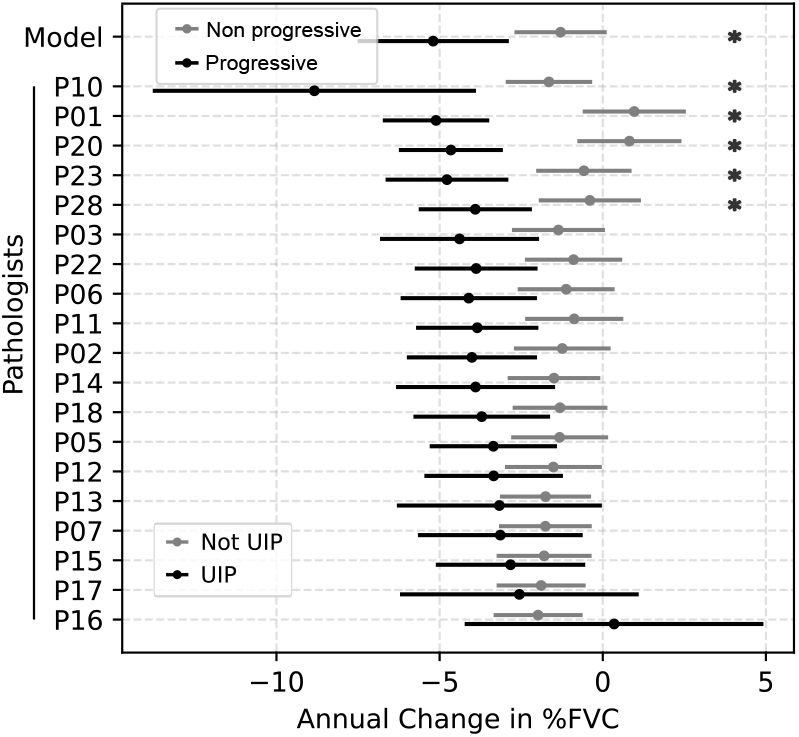
Comparison of prognostic performance between CRAI and pathologists using Generalized Linear Mixed Models. The forest plot demonstrates the estimated annual change in %FVC for different diagnostic categories in the TGH-TBLC cohort. The topmost row shows CRAI’s predictions, which classifies each case as progressive or non-progressive. The subsequent rows display the predictions based on 19 pathologists’ diagnoses, comparing UIP versus non-UIP classifications. The *x*-axis represents the estimated annual change in %FVC, with negative values indicating decline. Points and horizontal lines represent coefficient estimates and their 95% confidence intervals, respectively. CRAI demonstrated separation between non-progressive versus progressive groups without overlapping confidence intervals, indicating statistical significance. Among the 19 pathologists, only five achieved non-overlapping confidence intervals between their UIP and non-UIP diagnoses. These results suggest that CRAI’s prognostic performance surpasses that of most pathologists in predicting functional decline. The model was adjusted for age, sex, and baseline %FVC. FVC=forced vital capacity, UIP=usual interstitial pneumonia.

In the pathological diagnosis of interstitial pneumonia, the histopathological indicator for predicting patient survival prognosis is the presence or absence of UIP. Therefore, we compared the predictions of CRAI with each pathologist’s UIP diagnosis from the perspective of annual Δ%FVC. In the TGH-TBLC cohort, only five out of 19 pathologists could differentiate respiratory function prognosis (Figure 3). The model’s performance exceeded the prognostic prediction accuracy of many pathologists specializing in interstitial pneumonia. The interobserver agreement of UIP judgement among 19 pathologists were considered as low (Fleiss *κ* = 0.293).

### 3.2 Comparison of UIP Diagnosis and CRAI

We analyzed the prognosis predictions of CRAI and the diagnostic tendencies of pathologists for UIP. Although there were significant differences in UIP diagnostic tendencies between pathologists, it was found that most pathologists’ UIP diagnoses were similar to those of the model. We investigated the distribution of the cases using UMAP analysis based on whole slide level features. The cases classified as progressive group by CRAI corresponded with the cases diagnosed as UIP by many pathologists (Figure 4).

**Figure 4:**
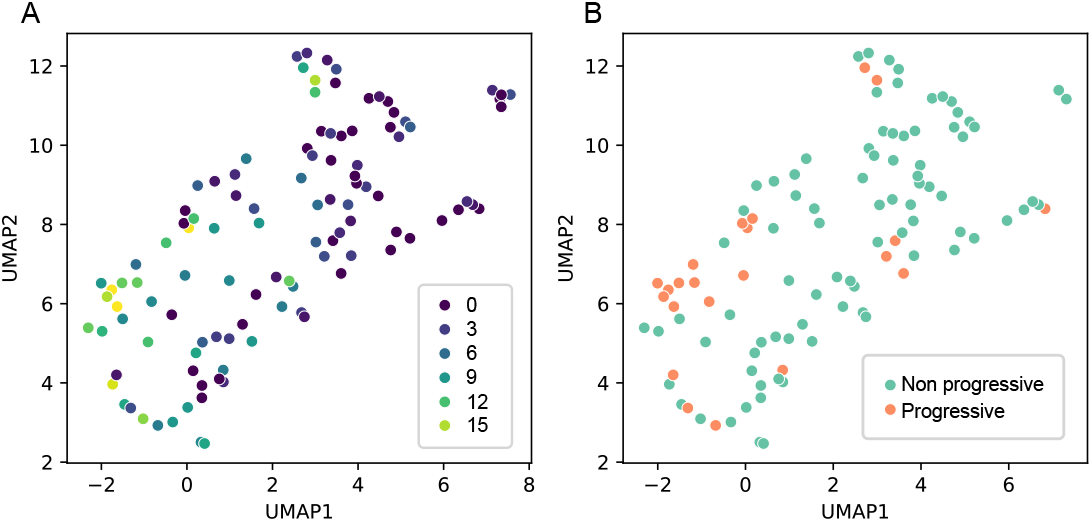
UMAP embedding of cases based on whole-slide level features. Each dot represents a single case, colored according to the number of pathologists who diagnosed UIP (A) and the classification by CRAI (B). Cases classified as progressiven disease correlate with those diagnosed as UIP by the majority of pathologists. UIP=usual interstitial pneumonia

### 3.3 Predictive Performance for Survival Prognosis

In the HRH-survival cohort, the median duration of follow-up was 1,561 days. At the last follow-up, 15 (31.2%) patients had died. The model predicted 38 cases as non-progressive and 10 cases as a progressive group, out of a total of 48 cases. In the non-progressive group, seven patients died, whereas in the progressive group, eight patients died. The group classified as progressive disease had a statistically significantly shorter survival time compared to the group inferred as a non-progressive group (Figure 5) (*p* = 0.034).

**Figure 5:**
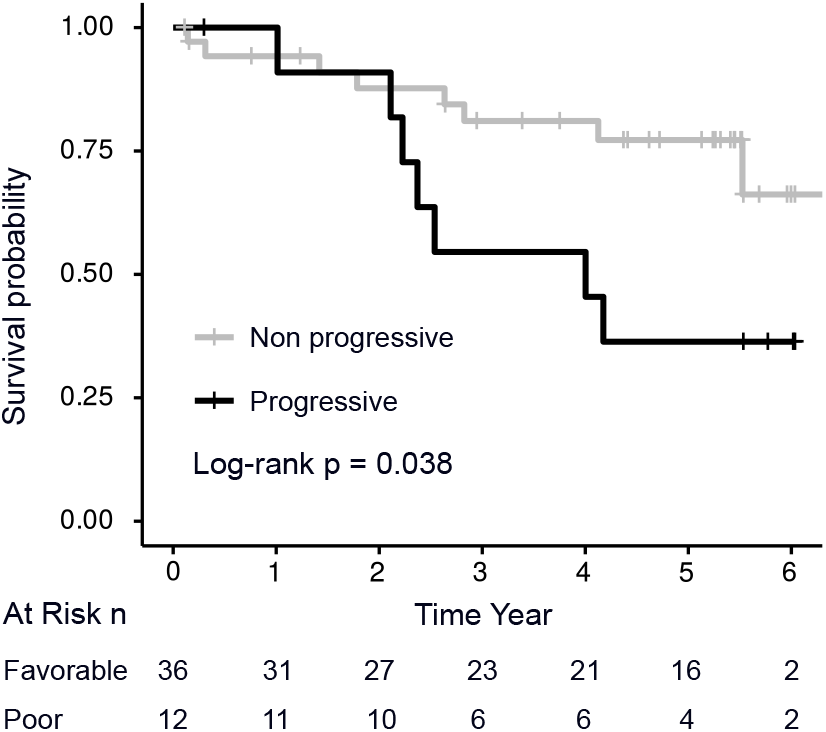
Kaplan-Meier Survival Curves for Progression Classification in the External Cohort. The progressive group as predicted by CRAI showed significantly shorter survival times compared to the non-progressive group (log-rank test, *p* = 0.038). The risk table below the graph shows the number of patients at risk in each group over time. These results demonstrate the model’s ability to effectively stratify patients based on their prognosis in an independent cohort from a different institution. Survival analysis was conducted on the HRH-survival cohort.

### 3.4 Pathological Characteristics of Cases Classified as Progressive Group by CRAI

SHAP (SHapley Additive exPlanations) analysis indicated that cases classified as progressive by the model showed a correlation between the presence of elastic fibers or nodular collagen deposition and the progressive group. Additionally, a correlation was observed between lymphoid follicles or FP/OP and the non-progressive group (Figure S2).

### 3.5 Validation of the AI model in an independent cross-domain cohort

Of 23 progressive cases, 18 (78%) were correctly predicted as progression by the AI model, while 10 of 19 non-progressive cases (53%) were correctly classified as non-progressive. The resulting odds ratio for disease progression was 4.0 (95% CI: 1.05–15.25; Fisher’s exact test *p* = 0.049), indicating that cases predicted as progressive by the model were approximately four times more likely to show clinical progression. Receiver operating characteristic analysis showed moderate discriminative performance, with an area under the curve (AUC) of 0.65. The overall classification performance yielded a sensitivity of 0.78, specificity of 0.53, and overall accuracy of 0.67.

## 4 Discussion

We have developed an explainable AI models, CRAI, the first AI model specifically designed to predict disease progression from TBLC specimens. The model offers comprehensive pathological analysis, and the results suggest its utility as a prognostic predictor. Notably, the model achieves considerable performance in differentiating pulmonary function deterioration. This result is particularly impressive considering that prognosis-predictive diagnosis of UIP has been challenging for most pathologists, even those experienced in ILD diagnosis. Low agreement of UIP diagnosis is confirmed by the low kappa coefficient, *κ* = 0.293 by the 19 pathologists in the TGH-TBLC cohort. The results suggest that, in many cases, the model could outperform prediction of deteriorating respiratory function compared to histological UIP diagnosis given by expert pulmonary pathologists.

The definition of short-term or long-term prognosis for ILD remains controversial. While the criteria for PPF are well-accepted, the setting of the observation period and the threshold for assessing respiratory function are arbitrary, and depending on the observation term, cases are categorized differently as seen in our TGH cases (Table 1). In this context, respiratory function serves as a well-known and independent surrogate marker for patient survival.^30, 31^ Due to the ease of data acquisition, we adopted it as the primary feature for establishing ground truth labels in the training data, while also utilizing it to compare the AI model’s and pathologists’ diagnoses.

However, given the significant variability in PFT results, we also prepared survival time as a more robust outcome for an external cohort. Consequently, in the external cohort, there was no significant difference in predicted %FVC between the progressive and non-progressive groups, but the estimated annual %FVC changes closely resembled the cross-validation results. In the survival analysis of the external validation cohort, the group predicted as a progressive group exhibited significantly shorter survival times, supporting the model’s robustness.

The cases predicted by CRAI as a progressive group generally possessed the features recognized by pathologists as UIP, aligning with the known association between UIP and progressive disease.^9, 12^ Interestingly, SHAP analysis indicated that nodular collagen fiber deposition and elastosis were progression factors, while fibroplasia (FP/OP) and lymphoid follicles were factors associated with non-progressive group, demonstrating similarity with the diagnostic process of UIP by pathologists.

Importantly, CRAI maintained a significant association with disease progression when validated against an external dataset, representing a distinct imaging domain, including differences in scanner hardware, institutional staining protocols, and slide preparation processes. Such heterogeneity is a major challenge for AI models in digital pathology due to domain shift. The consistent predictive association observed in this study suggests that the model relies on biologically meaningful morphological patterns rather than scanner-specific artifacts, highlighting its potential robustness for real-world clinical implementation.

Instead of adopting an end-to-end model using recent large foundation models that directly predicts outcomes such as respiratory function prognosis or survival prognosis, CRAI emphasized extracting pathologically interpretable findings. This approach is particularly suitable in the context of ILD diagnosis, which differs qualitatively from tumor diagnosis. While tumors exhibit significant variations in size, shape, and surrounding microenvironments, ILD presents as a mixture of inflammatory and post-inflammatory reactions composed of normal cellular components. It is also important for cases requiring explainability and for those with a limited amount of digital data compared to common neoplastic diseases. While pathologists can easily recognize findings such as fibrosis or inflammation, the lack of standardized quantification methods for these features has led to variability in final diagnoses. These detected findings are not only utilized for the specific downstream task such as the patient’s prognosis but also serve as a platform for various histopathological analyses.

Compared to our previous MIXTURE model targeting SLB,^15^ this model incorporates several improvements considering the histological characteristics of TBLC. One key difference is the sampling site. In SLB, the peripheral parts of the lung are sampled, and large bronchovascular bundles are less likely to be included. In contrast, TBLC specimens are collected transbronchially, inevitably including bronchi and surrounding tissues as a major part of biopsy sample. Since diagnostically important lesions are primarily located in the lung parenchyma rather than in the bronchovascular bundles, and SLB samples are large enough to minimize the influence of bronchovascular structures, however; TBLC tissues are comparatively small, making the presence of bronchovascular bundles more impactful within the same tissue. Therefore, separating these areas from evaluation in the AI model is crucial.

This study has limitations. A major issue is the small number of cases available for consideration to cover disease progression. TBLC is a relatively recently introduced technique, and in some regions, only a limited number of institutes can perform. In particular, it is challenging to collect cases with long-term follow-up necessary to evaluate true disease progressive behavior with such a new technique. However, in places such as Japan and Europe, TBLC is increasingly being adopted by more facilities due to its less invasive nature. As more cases accumulate in the future, further improvement in the model’s performance and validation will become possible.

The current model focuses solely on predicting prognosis from the histological findings of TBLC. With this approach, it may be possible to identify potentially progressive cases without observing them over a certain period, as per the criteria for PPF. It could detect progressive cases at the stage of ILA as our data indicated, which are often incidentally discovered in asymptomatic individuals during screening.^29^ Furthermore, by combining multiple modalities such as clinical data, physiological measurements, and radiological imaging, it could enable more precise identification of progressive patients than is currently possible, leading to more personalized and timely therapeutic interventions as our previous work.^32^

In summary, we have developed CRAI, the first deep learning-based model to comprehensively analyze ILD TBLC tissues. This model enables stratification of disease progression and will not only provide a foundation for objective tissue analysis of ILD but also lead to standardized diagnosis to optimize patient care.

## Data Availability

Upon approval from the Institutional Review Board at Nagasaki University Hospital, a subset of individual participant data can be shared for research purposes upon reasonable request to the corresponding author.

https://github.com/FukuokaLab/CRAI

## 5 Contributors

W.U. and J.F. conceived the project. K.K., Yu.Ki., Ya.Koh., Y.S., and Ya.Kon. contributed clinical cases and related information. Under the supervision of J.F., L.C., and H.S., W.U. trained the models, analyzed the experimental data, and performed statistical analysis. M.O., Y.N., K.L., the CRYOSOLUTION study group, and J.F. conducted the diagnostic assessments. J.F. and Ya.Kon. validated the results. W.U. drafted the original manuscript, which was subsequently revised by E.O., K.L., and J.F. All authors reviewed and approved the final manuscript. J.F. acquired funding for the study.

## 6 Data Sharing

Upon approval from the Institutional Review Board at Nagasaki University Hospital, a subset of individual participant data can be shared for research purposes upon reasonable request to the corresponding author. Our experiments utilized the open-source code of YoLoV7 (https://github.com/WongKinYiu/yolov7). The scripts for the CRAI system are available (https://github.com/FukuokaLab/CRAI).

## 7 Acknowledgement

This paper is based on results obtained from a project, JPNP20006, commissioned by the New Energy and Industrial Technology Development Organization (NEDO) and partly supported by the research group of diffuse lung disease under the Japanese Ministry of Health, Labor, and Welfare (20FC1033).

## 8 Declaration of generative AI and AI-assisted technologies in the manuscript preparation process

During the preparation of this work the author(s) used Claude-3.5-Haiku (version 20241022) in order to proofread the language. After using this tool/service, the author(s) reviewed and edited the content as needed and take(s) full responsibility for the content of the published article.

## Appendix A The definition of slide-level findings

1. **Lung**: The area of lung parenchyma, measured in square millimeters (*mm*^2^).
2. **Lesion**: The proportion of the lesion (non-normal component) within the lung area, calculated using the MIXTURE 5× model.
3. **Fibrotic/Cellular Ratio**: The ratio of the fibrotic area to the cellular area within the lung area, calculated using the MIXTURE 5× model. Regions classified as lymphoid follicles by the MIXTURE 5× model were included in the cellular area.
4. **Lymphoid Follicles**: The number of lymphoid follicles within the lung area, calculated using the MIXTURE 5× model. Adjacent patches identified as lymphoid follicles were considered a single lymphoid follicle.
5. **Dense Fibrosis**: The area of dense fibrosis within the lung area, calculated using the MIXTURE 20× model.
6. **Bronchial Epithelium**: The area of bronchial epithelium within the lung area, derived from the MIXTURE 20× model.
7. **FP/OP**: The number of fibroblastic proliferation (FP) or organizing pneumonia (OP). These two findings are difficult to differentiate, especially in high-power field views. The “immature fibrosis” area was detected using the MIXTURE 20× model, and the number of independent immature fibrosis regions within the lung area was counted.
8. **Max Collagen Around FP/OP**: The maximum collagen index surrounding the FP/OP detected above.
9. **Lymphocyte Density in Dense Fibrosis**: The number of lymphocytes per square millimeter (*mm*^2^) in the dense fibrosis area.
10. **Lymphocyte Density in Non-Dense Fibrosis**: The number of lymphocytes per square millimeter (*mm*^2^) in the non-dense fibrosis area.
11. **Collagen in Non-Dense Fibrosis**: The mean collagen index in the non-dense fibrosis area within the lung.
12. **Collagen Edge**: The amount of collagen edge, capturing abrupt changes in fibrosis. Collagen is derived from the Collagen model. The edge is detected by a Sobel filter, subtracting edges related to tissue boundaries.
13. **Collagen Edge/Dense Fibrosis**: The ratio of the collagen edge to dense fibrosis.
14. **Collagen Forming the Blob**: Proportion of collagen forming nodular clusters. Collagen is derived from the Collagen model. Blobs are detected by Laplacian of Gaussian. The proportion of collagen within the blob to the total amount of collagen is calculated.
15. **Elastic Fiber**: The mean elastic fiber index in lung area.
16. **Diffuse**: Proportion of diffuse features in the lung area, identified by the 1× model.
17. **Patchy**: Proportion of patchy features in the lung area, identified by the 1× model.
18. **Honeycomb**: Proportion of honeycomb features in the lung area, identified by the 1× model.

## Appendix B Tables

**Table S1:**
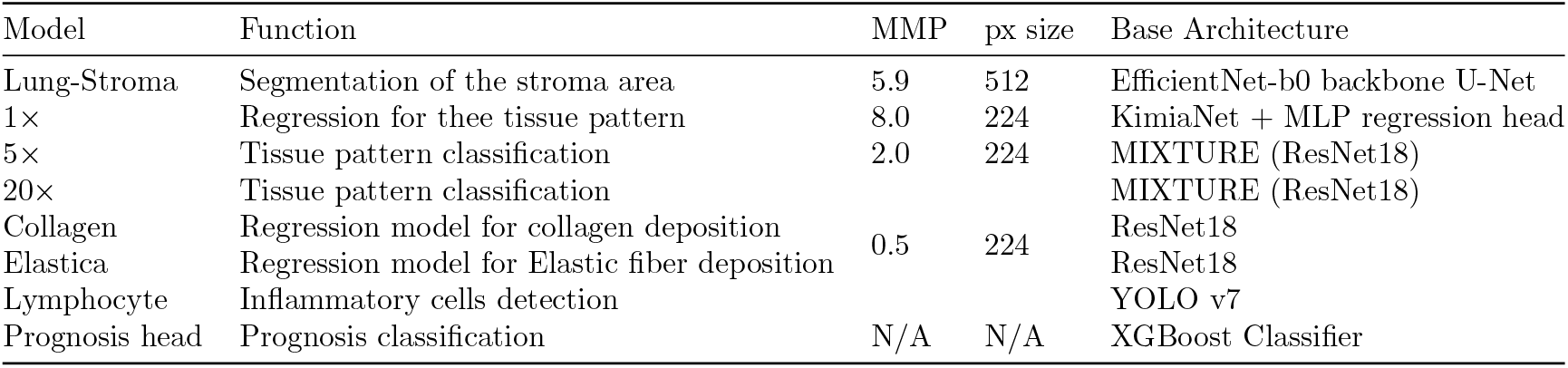
Description of Model Architectures. MMP=micron per pixel

**Table S2:** The correspondance between MIXTURE and new model (CRAI) classification (5×)

| **MIXTURE** | **CRAI** |
| --- | --- |
| Complete Normal | Normal |
| Acellular fibrosis | Fibrotic area |
| Cellular and fibrotic IP |  |
| Pale tissue |  |
| NSIP/Cellular IP | Cellular area |
| Lymphoid follicle | Lymphoid follicle |
| Edge | Other |
| Other |  |

**Table S3:** The correspondance between MIXTURE and new model (CRAI) classification (20×)

| **MIXTURE** | **CRAI** |
| --- | --- |
| Dense fibrosis | Fibrosis |
| Elastosis |  |
| Fibroblastic foci | FP/OP |
| Mucin | Mucin |
| Bronchiolar epithelium | Bronchiolar epithelium |
| Fat | Other |
| Lymphocytes aggregation |  |
| Other |  |

**Table S4:** Coefficient Estimates for Annual %FVC Change from Linear Mixed Models: Comparison between CRAI (non progressive/progressive) and Pathologists’ (UIP/non-UIP) Diagnostic Classifications in TGH-TBLC cohort. UIP=usual interstitial pneumonia, CI=confidence interval

| Model | Non-progressive |  | Progressive |  | Age |  | Male Sex |  |
| --- | --- | --- | --- | --- | --- | --- | --- | --- |
|  | Coef. | 95% CI | Coef. | 95% CI | Coef. | 95% CI | Coef. | 95% CI |
| Model | -1.293 | (-2.707, 0.121) | -5.198 | (-7.515, -2.880) | -0.149 | (-0.329, 0.030) | 1.043 | (-2.481, 4.567) |

| Pathologists | not UIP |  | UIP |  | Age |  | Male Sex |  |
| --- | --- | --- | --- | --- | --- | --- | --- | --- |
|  | Coef. | 95% CI | Coef. | 95% CI | Coef. | 95% CI | Coef. | 95% CI |
| P01 | 0.965 | (-0.615, 2.546) | -5.110 | (-6.740, -3.480) | -0.070 | (-0.233, 0.093) | 1.683 | (-1.565, 4.931) |
| P02 | -1.240 | (-2.721, 0.240) | -4.010 | (-6.005, -2.014) | -0.124 | (-0.306, 0.058) | 0.639 | (-3.037, 4.316) |
| P03 | -1.359 | (-2.783, 0.065) | -4.390 | (-6.827, -1.954) | -0.164 | (-0.346, 0.019) | 0.598 | (-3.012, 4.209) |
| P05 | -1.322 | (-2.808, 0.164) | -3.353 | (-5.304, -1.403) | -0.191 | (-0.367, -0.016) | 0.339 | (-3.149, 3.826) |
| P06 | -1.122 | (-2.607, 0.363) | -4.107 | (-6.194, -2.019) | -0.193 | (-0.379, -0.008) | 0.780 | (-2.826, 4.386) |
| P07 | -1.759 | (-3.179, -0.339) | -3.140 | (-5.663, -0.617) | -0.120 | (-0.304, 0.063) | 0.503 | (-3.192, 4.198) |
| P10 | -1.650 | (-2.975, -0.325) | -8.839 | (-13.792, -3.886) | -0.142 | (-0.325, 0.040) | 1.393 | (-2.246, 5.032) |
| P11 | -0.875 | (-2.379, 0.629) | -3.847 | (-5.721, -1.973) | -0.145 | (-0.325, 0.034) | 0.958 | (-2.647, 4.563) |
| P12 | -1.514 | (-2.998, -0.030) | -3.344 | (-5.466, -1.222) | -0.127 | (-0.314, 0.059) | 0.923 | (-2.826, 4.673) |
| P13 | -1.754 | (-3.150, -0.359) | -3.169 | (-6.309, -0.029) | -0.144 | (-0.330, 0.042) | 0.862 | (-2.855, 4.579) |
| P14 | -1.494 | (-2.914, -0.074) | -3.900 | (-6.337, -1.463) | -0.150 | (-0.331, 0.030) | 0.758 | (-2.842, 4.357) |
| P15 | -1.797 | (-3.251, -0.343) | -2.826 | (-5.116, -0.537) | -0.122 | (-0.301, 0.058) | 1.351 | (-2.269, 4.970) |
| P16 | -1.981 | (-3.345, -0.618) | 0.347 | (-4.231, 4.924) | -0.152 | (-0.336, 0.033) | 0.953 | (-2.729, 4.636) |
| P17 | -1.888 | (-3.252, -0.524) | -2.556 | (-6.214, 1.102) | -0.143 | (-0.327, 0.041) | 0.667 | (-3.000, 4.333) |
| P18 | -1.310 | (-2.762, 0.142) | -3.708 | (-5.801, -1.616) | -0.120 | (-0.302, 0.062) | 1.093 | (-2.536, 4.722) |
| P20 | 0.817 | (-0.780, 2.413) | -4.655 | (-6.249, -3.062) | -0.072 | (-0.240, 0.096) | 0.553 | (-2.769, 3.876) |
| P22 | -0.894 | (-2.384, 0.597) | -3.881 | (-5.761, -2.000) | -0.114 | (-0.298, 0.071) | 0.953 | (-2.716, 4.621) |
| P23 | -0.578 | (-2.039, 0.882) | -4.775 | (-6.656, -2.894) | -0.116 | (-0.293, 0.061) | 0.602 | (-2.913, 4.117) |
| P28 | -0.396 | (-1.962, 1.169) | -3.907 | (-5.640, -2.173) | -0.133 | (-0.312, 0.047) | 1.087 | (-2.512, 4.685) |

**Table S5:** Coefficient Estimates for Annual %FVC Change from Linear Mixed Models in ILA subgroup in TGH-TBLC cohort. CI=confidence interval

| Model | Non progressive |  | Progressive |  | Age |  | Male Sex |  |
| --- | --- | --- | --- | --- | --- | --- | --- | --- |
|  | Coef. | 95% CI | Coef. | 95% CI | Coef. | 95% CI | Coef. | 95% CI |
| Model | -2.853 | (-7.090, 1.384) | -8.093 | (-13.766, -2.420) | -0.244 | (-0.892, 0.405) | -3.256 | (-14.236, 7.724) |

**Table S6:**
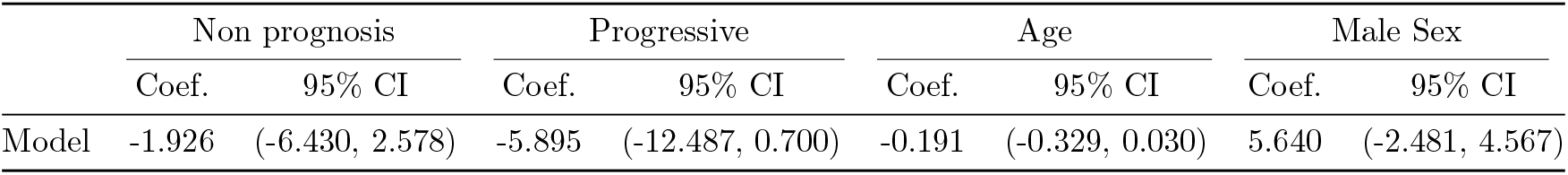
Coefficient Estimates for Annual %FVC Change from Linear Mixed Models in HRH-PFT (external) cohort. CI=confidence interval

| Model | Non prognosis |  | Progressive |  | Age |  | Male Sex |  |
| --- | --- | --- | --- | --- | --- | --- | --- | --- |
|  | Coef. | 95% CI | Coef. | 95% CI | Coef. | 95% CI | Coef. | 95% CI |
| Model | -1.926 | (-6.430, 2.578) | -5.895 | (-12.487, 0.700) | -0.191 | (-0.329, 0.030) | 5.640 | (-2.481, 4.567) |

## Appendix C Figures

**Figure S1:**
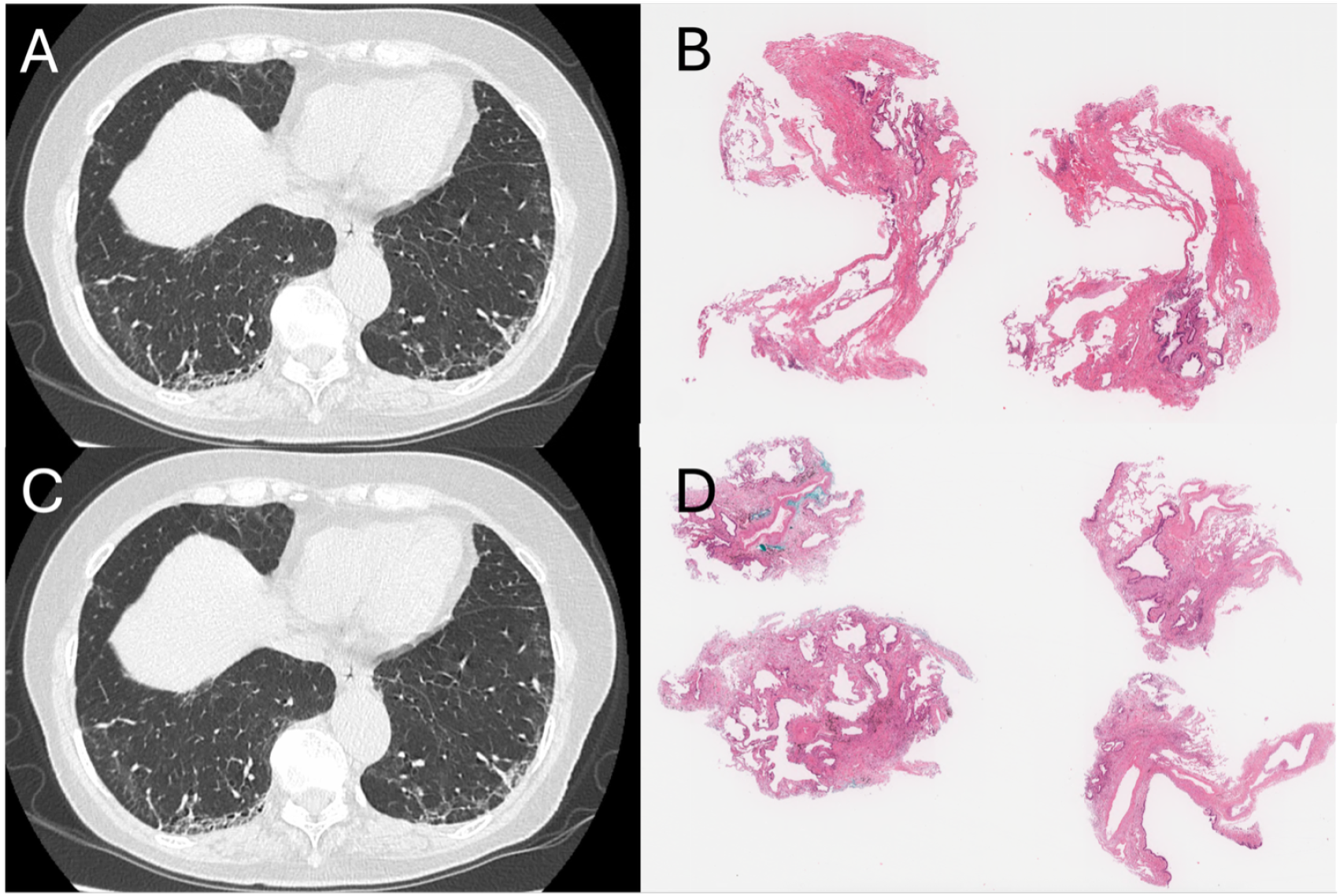
Two representative cases of interstitial lung abnormalities (ILA). A, B: The baseline %FVC was 132.1%, but it progressed to 133.5% after one year and 123.1% after two years, indicating clinical progression. C, D: The baseline %FVC was 123.6%, but it progressed to 113.0% after one year and 95.6% after two years, also indicating clinical progression. Pathologically, both cases exhibited a usual interstitial pneumonia (UIP) pattern, and the model predicted a “progressive” for them.

**Figure S2:**
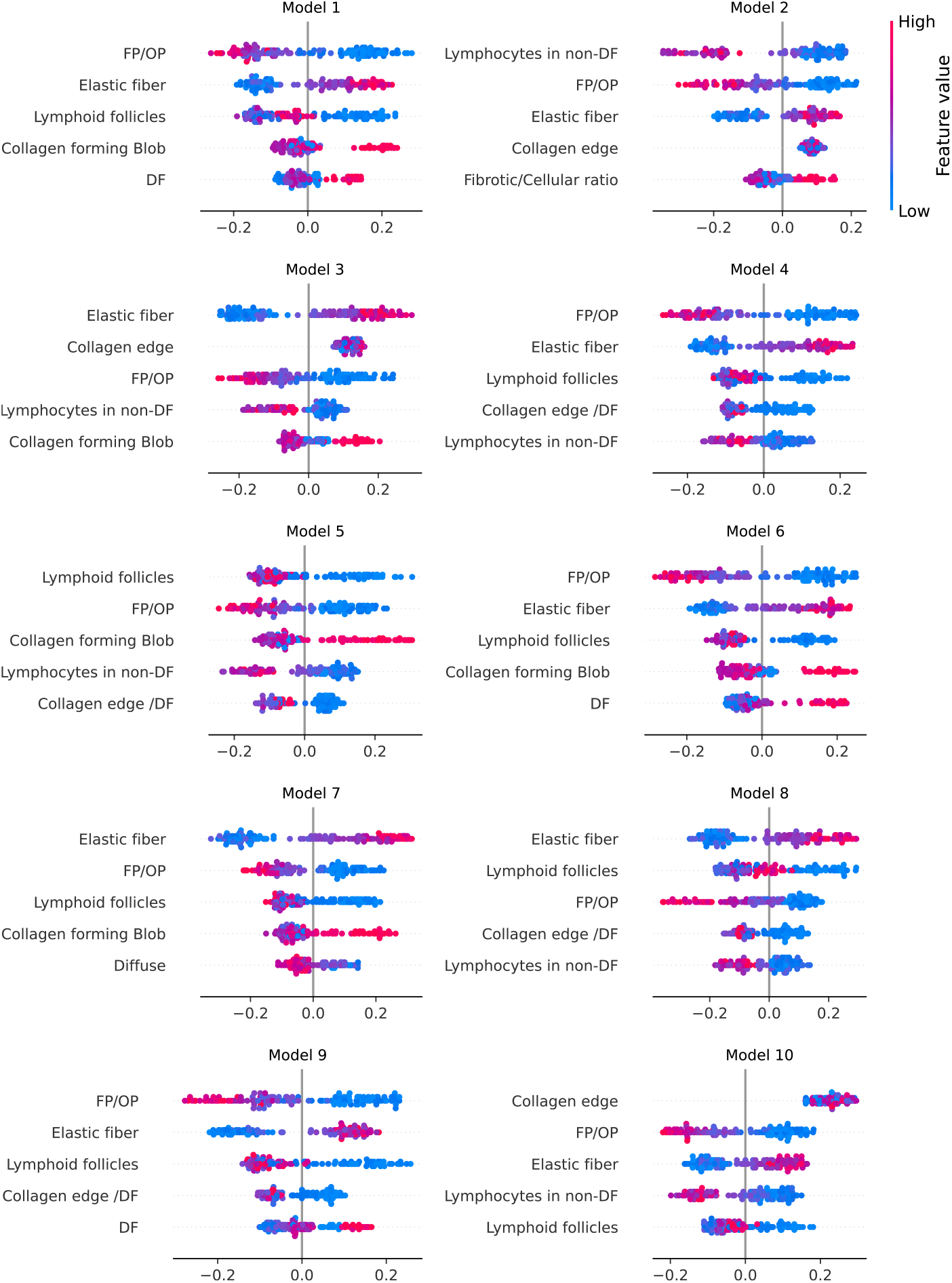
SHAP (SHapley Additive exPlanations) summary plots for ten XGBoost models. For each model, the top five most influential findings are displayed based on their mean absolute SHAP values. Red points indicate positive correlation with progressive disease, while blue points indicate negative correlation. The horizontal location shows the SHAP value, representing the impact of each finding on the model output. Notably, across multiple models, “elastic fiber” consistently demonstrates strong positive correlation with “progressive”, whereas “FP/OP” shows strong negative correlation, suggesting its association with preferable prognosis. The distribution of points along the x-axis represents the magnitude of each finding’s impact on the model predictions.

**Figure S3:**
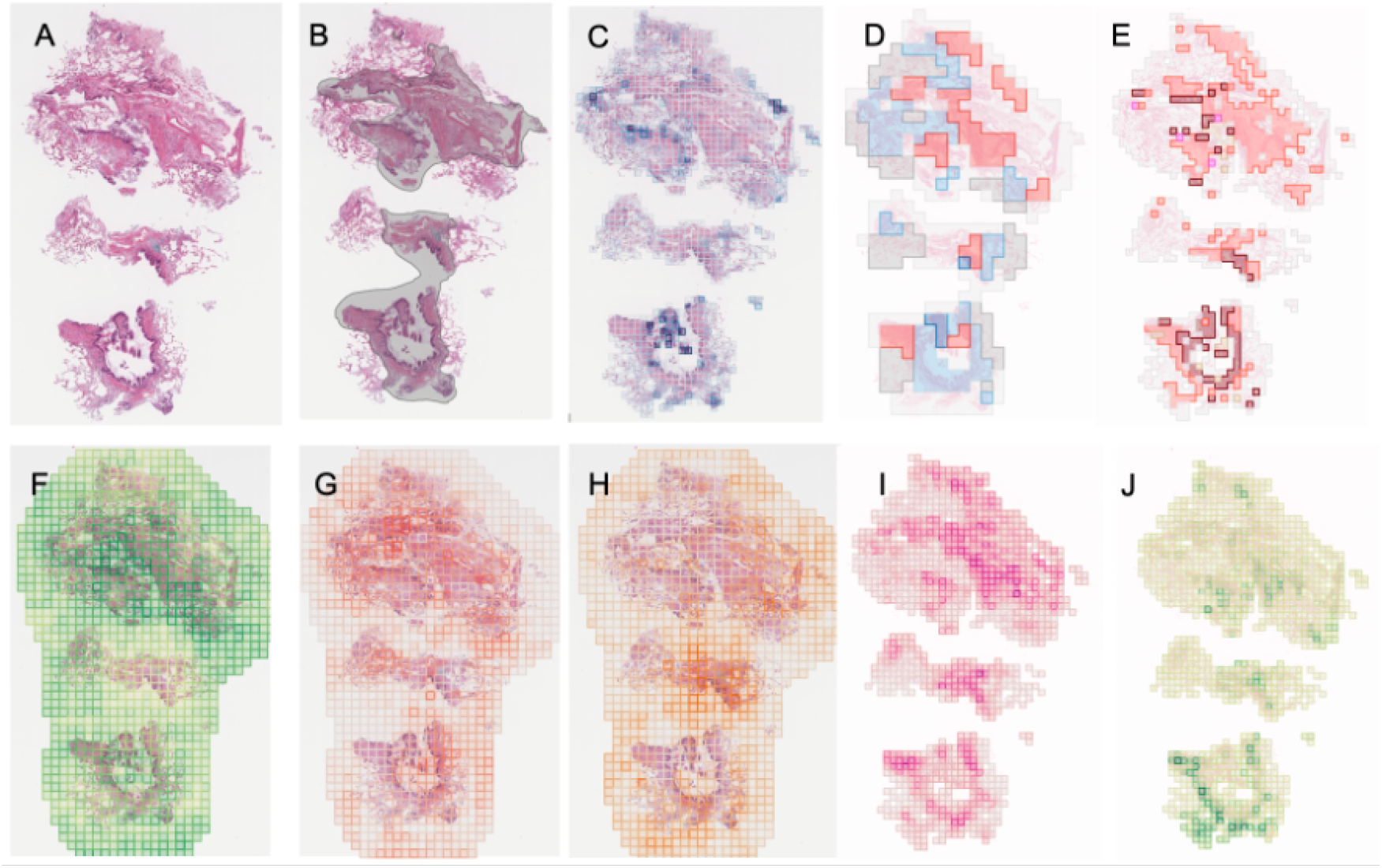
Representative results of CRAI. (A) Original H&E-stained whole slide image. (B) Lung-stroma model highlighting broncovascular bundles in gray. (C) Spatial distribution of lymphocyte density as predicted by the lymphocyte model. (D) Classification results of the 5× model, where *fibrotic areas* are shown in red, *cellular areas* in light blue, *lymphoid follicles* in blue (not present in this case), *normal tissue* in gray, and *other findings* in white. (E) Results of the 20× model, with *fibrosis* marked in orange, *bronchiolar epithelium* in brown, *FP/OP pattern* in pink, *mucin* in yellow, and *other findings* in gray. (F-H) Predictions of three separate 1× models, showing the distribution of *diffuse, patchy*, and *honeycomb patterns*, respectively. (I) Predicted distribution of *collagen fibers* by the collagen model. (J) Predicted distribution of *elastic fibers* by the elastica model.

## Notes

### Author Declarations

Ethics Committee of Nagasaki University Hospital gave ethical approval for this work

### Summary of Updates

Section 2.15 and 3.5 added (external validation)

